# Development of a Model for Differentiating PDAC from Benign Pancreatic Conditions: A Prospective Case-control Study

**DOI:** 10.1101/2022.03.23.22272803

**Authors:** Mohamed Zardab, Vickna Balarajah, Abhirup Banerjee, Konstantinos Stasinos, Amina Saad, Ahmet Imrali, Christine Hughes, Rhiannon Roberts, Ajith Vajrala, Claude Chelala, Hemant M Kocher, Abu Z M Dayem Ullah

**Affiliations:** Centre for Tumour Biology, Barts Cancer Institute, Queen Mary University of London, London, UK; Barts and the London HPB Centre, The Royal London Hospital, Barts Health NHS Trust, London, UK; Barts Pancreas Tissue Bank, Barts Cancer Institute, Queen Mary University of London, London, UK; Centre for Cancer Biomarkers and Biotherapeutics, Barts Cancer Institute, Queen Mary University of London, London, UK

**Keywords:** PDAC, symptoms, blood test, comorbidity, prediction algorithm

## Abstract

**Background & aims:** Pancreatic ductal adenocarcinoma (PDAC) continues to be a devastating disease with late diagnosis and poor overall survival, complicated by clinical presentations similar to benign pancreatic diseases. We aimed to analyse clinical parameters with the goal of developing a prediction model for differentiating suspected PDAC from benign pancreatic conditions.

**Methods and results:** We used a prospectively recruited cohort of patients with pancreatic disease (n=762) enrolled at the Barts Pancreas Tissue Bank between January 1, 2008 and September 21, 2021 to perform a case-control study examining the association of PDAC (n=340) with predictor variables including demographics, comorbidities, lifestyle factors, presenting symptoms and commonly performed blood tests. Using a machine learning approach, candidate PDAC risk-prediction algorithms were trained on 75% of the cohort, using a subset of the predictor variables identified from a preliminary observational association study. Models were assessed on the remaining 25%. Multiple imputed datasets were used for both training and validation to accommodate for unknown data.

Age (over 55), weight loss in hypertensive patients, recent symptom of jaundice, high serum bilirubin, low serum creatinine, high serum alkaline phosphatase, low lymphocyte count and low serum sodium were the most important features when separating putative PDAC cases from less severe pancreatic conditions. A simple logistic regression model had the best performance with an area under the curve (AUC) of 0.88. Setting a probability threshold of 0.17 guided by the maximum *F*_*2*_ score, a sensitivity of 95.6% was reached in the full cohort which could lead to early detection of around 84% of the PDAC patients.

**Conclusion:** The resultant prediction model significantly outperformed the current UK guidelines for suspected pancreatic cancer referral and could improve detection rates of PDAC in the community. After further work this approach could lead to an easy to understand, utilisable risk score to be applied in the primary and secondary care setting for referring patients to specialist hepato-pancreatico-biliary services.

## Introduction

Pancreatic ductal adenocarcinoma (PDAC) is an aggressive malignant disease of the pancreas with a global annual death-toll of over 400000 and overall survival between 2-7%.^1^ It continues to be a diagnostic and therapeutic challenge with little survival improvement over the decades.^2^ PDAC is associated with poor prognosis due to several factors.^3^ Low incidence (∼12 per 100,000),^4^ non-specific symptoms, late presentation, aggressive and resistant tumour biology with early distant metastasis, and lack of specific and sensitive biomarkers or imaging of early disease all contribute to its high mortality. The only possible curative option is surgical resection with adjuvant therapy offering 5-year overall survival rates between 15-25% (>40% overall survival in selected subgroups).^2,5^ Unfortunately, ∼80% patients present with unresectable disease,^2^ which makes PDAC a disease with drastically worse morbidity and mortality than benign pancreatic pathologies^6^ and other non-PDAC pancreatic cancers.^7,8^

Prospective and retrospective studies of patients in the community have shown a low prevalence and predictive value of clinical symptoms,^9^ although jaundice and weight loss have been shown to have a strong association with PDAC in clinical practice.^10^ While serum carbohydrate antigen 19-9 (CA19-9) is widely used for monitoring PDAC treatment response and recurrence, its utility as diagnostic biomarker for screening in the general population is limited due to low sensitivity and specificity.^11^ New-onset diabetes has recently been considered as a significant indicator of PDAC in the older population (>60 years of age) and screening clinical trials are underway for this patient subgroup.^11–13^ Symptom-based pathways to diagnosis in primary care setting is also complicated by comorbidities,^16^ and patients are delayed for referral to specialist consultation at secondary/tertiary care even when presenting with clinically associated symptoms such as back pain, diabetes and weight loss.^14^ On the other hand, even if the referrals are followed in accordance to national guidelines, chances of detecting early, potentially curable disease is miniscule, since most patients in this clinical pathway are presented with metastatic disease.^15^

Predictive algorithms for identifying PDAC risk groups have been developed from large primary care databases,^16,17^ mainly focusing on presenting symptoms and demographic characteristics. Results from such symptom-based cancer decision support tools (CDSTs) suggest improved discriminatory ability;^17,21^ however they are over-fitted for certain patient groups and require continuous refinement with inclusion of additional features.^16,18^ Pre-existing medical conditions are rarely considered in these algorithms and so are the associated commonly performed laboratory tests, which could enhance the performance. Data source can also be a factor as pre-diagnostic symptom spectrum appears to be different in studies conducted using secondary/tertiary care than primary care data.^19^ Since benign pancreatic conditions such as chronic pancreatitis are known risk factors and demonstrate almost identical clinical presentations to PDAC,^18,19^ differentiation of PDAC from benign conditions within CDST workflow can also be a useful but challenging addition for reducing false positive detection.

Digital technologies would enable future healthcare professionals to take advantage of automated referral pathways, suggested by algorithms using routinely collected clinical data.^20^ This has become a realistic possibility with the advent of machine learning, which allows handling vast numbers of clinical variables from patients’ medical records and searching for non-linear and interactive combinations to predict target outcomes. In this study, towards the broader goal of developing a digital referral tool for suspected PDAC, we utilised medical histories from a prospective cohort of patients with various pancreatic diseases who were treated at a specialised hepato-pancreatico-biliary (HPB) clinic in the UK. We identified a compendium of discriminatory symptoms and commonly performed laboratory test results, appropriately adjusted for common confounding demographic and clinical factors, that could differentiate PDAC from benign pancreatic conditions. Then, we aimed to develop an optimal machine learning model for potential use in the primary care setting to guide referral decisions for suspected pancreatic conditions.

### Materials and methods Study setting

All data utilised for this research were collected from the Barts Pancreas Tissue Bank (BPTB; https://www.bartspancreastissuebank.org.uk/)) study, with written informed consent from patients recruited at Barts Health NHS Trust (BHNT) (Hampshire B Research Ethics Committee: 18/SC/0630). The BPTB is a repository of biospecimens and associated clinical data collected from patients (≥18 years) referred to the specialist clinic of HPB surgery at BHNT (Barts and the London HPB Centre) for confirmed or suspected malignant or benign diseases of the pancreas and other diseases of hepatobiliary origin. The biobank also recruits healthy volunteers to be used as contemporaneous comparison cohort.

Data on BPTB participants’ ‘health’ features are entered into a bespoke, encrypted, secure database using a predefined 49-item questionnaire, of which 37 items are completed by trained tissue collection officers in a face-to-face interview during recruitment.^21^ These include demographic information, anthropometric measurements, persistent symptoms, comorbidities, regular use of specific medication, lifestyle behaviours, family life, and family history of pancreatic and other cancers. Another 12 items of information (with varying granularity) about clinical diagnoses, treatment including surgical and non-surgical interventions, and test results including blood test, urine test, imaging, and histopathology reports are populated from their electronic health records (EHR) data at BHNT. Data is further cross-checked and verified by trained clinicians to ensure accuracy.

### Study design

A ‘nested’ case-control study was carried out to examine the association of PDAC diagnosis with a set of predictor variables, in comparison to patients with non-malignant pancreatic diseases (PnC), and subsequently to develop a risk-prediction algorithm that may separate diagnosis of PDAC from PnC. The choice of PnC as a reference group was driven by the perceived clinical utility of the potential findings. Patients with pancreatic diseases, malignant and non-malignant alike, are believed to demonstrate similar symptomatic presentation (e.g., jaundice) and biochemical profile (e.g., elevated CA19-9 or deranged Liver Function Tests),^19^ therefore identifying any distinctive clinical profile may have an impact on the evidence-based decision making of the referral and diagnostic pathway.

We extracted relevant clinical data for all patients registered with the BPTB study between January 1, 2008 and September 21, 2021 and diagnosed with any pancreatic condition (*N*=780 of 1371) by excluding healthy controls and those with non-pancreatic conditions (n=588). A diagnosis of pancreatic neoplasm is recorded in the BPTB database using the ICD-O-3 (International Classification of Diseases - Oncology, 2013) multi-axial classification for the site and the histology of the neoplasm; the non-neoplastic disorders of pancreas are coded using the ICD-10 codes (2019) (Supplementary Table 1). The final diagnosis recorded in the database is recorded using hierarchical ascertainment mechanisms: histology reports, multidisciplinary team meetings outcomes, consultation notes from the oncology or gastroenterology specialist, findings from test results (e.g., radiological and endoscopic reports) and/or commissioning data sets diagnosis entry. The date on the first confirmatory histology report is used as the date of diagnosis, otherwise the earliest of dates from the other evidences is used. Complex diagnoses or cases with incomplete data are agreed on by an adjudication group of clinicians (HMK, KS, MZ).

Patients were finally assigned to one of the three groups (in the order of priority) - PDAC, Other pancreatic cancers (PC), and PnC, based on their final diagnosis entry in the database. For each individual within a specific group, the date associated with the earliest diagnosis was considered as the index date. This is particularly applicable for individuals in the PnC group who can have final diagnoses of different pancreatic diseases. Care was taken to exclude patients, with secondary tumour to the pancreas (n=2), with concomitant primary cancer in another body site (n=6), and those who were assigned a provisional diagnosis but were not reassigned to a confirmed diagnosis (n=1) to avoid biases (Figure 1). Comorbidity and symptoms history were not collected for nine patients. Since the study focused solely on PDAC, we excluded patients with other pancreatic cancer (n=58) (Supplementary Table 2).

**Figure 1.**
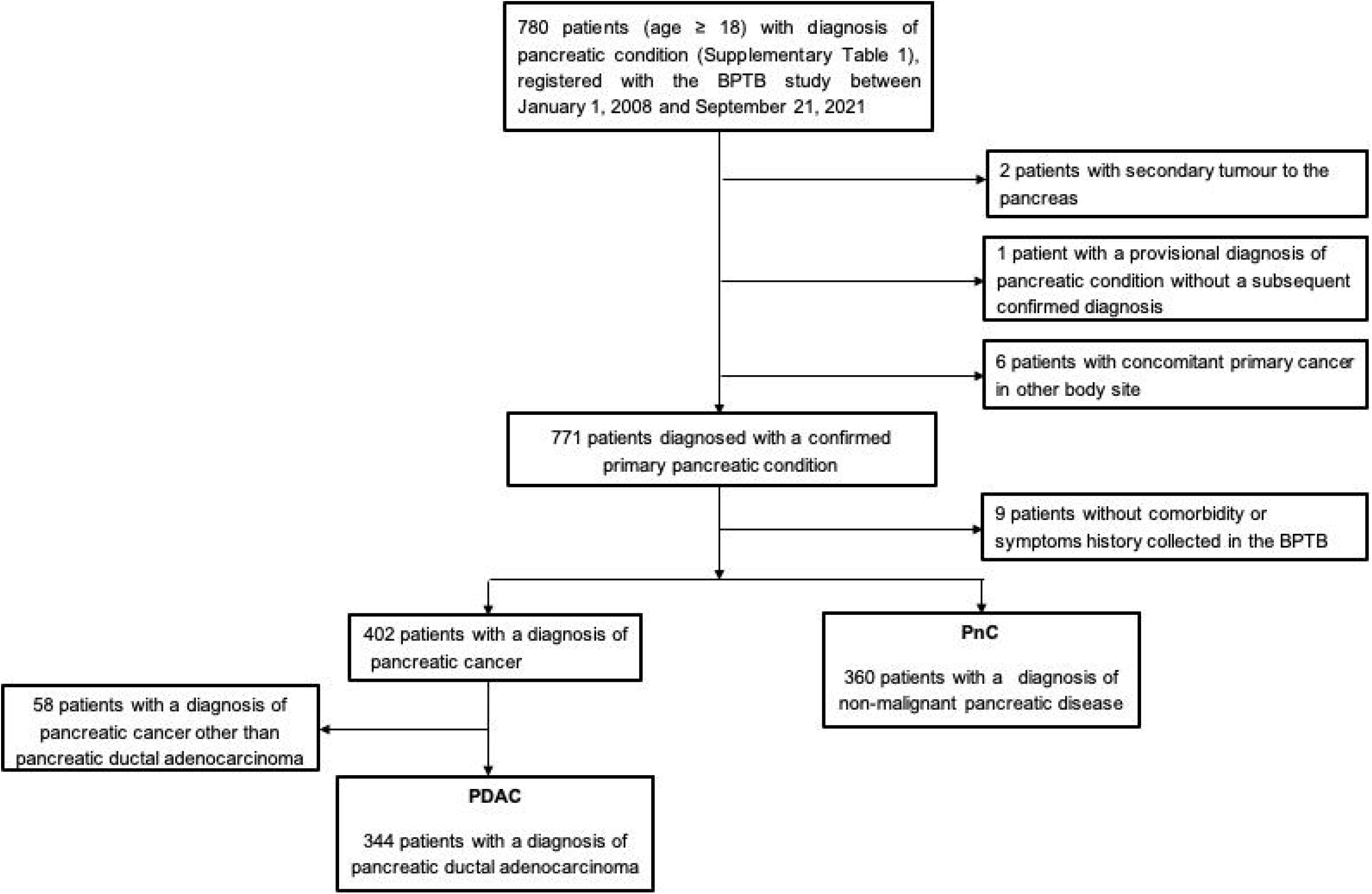
CONSORT diagram for selection of patients in the nested case-control study.

### Assessment of Predictor variables

Details on the predictor variables and their categorisation are provided in Supplementary Table 3. Demographic details included in the study were gender (Male, Female); age (<55, 55-64, 65-74, 75+); ethnic origin (Caucasian, Afro-Caribbean, South Asian, or Other). Lifestyle variables included smoking history of alcohol consumption (current, past, or never) and body mass index (BMI). Clinical variables relating to comorbidities (present or absent) comprised of; diabetes, hypertension, high cholesterol, chronic respiratory disease, cardiovascular disease, chronic kidney disease, liver disease and previous cancer diagnosis. Both lifestyle and comorbidity variables were recorded based on the patients’ status at the time of recruitment.

The following eleven symptoms that might herald a diagnosis of gastrointestinal problems were included as binary outcomes (present vs absent): pain, jaundice, weight loss, nausea, vomiting, diarrhoea, constipation, fatigue, loss of appetite, pruritus, and steatorrhea. The presence or absence of these symptoms within the one-year period before recruitment were confirmed by the TCOs in a face-to-face interview. A composite symptom was also derived in the form of change in bowel habits, manifested in any of diarrhoea, constipation, or steatorrhea.

Twenty-two laboratory tests, commonly requested by clinicians for suspected HPB problems, were examined (Supplementary Table 3). For each test, the instance closest to the recruitment date and conducted within six months before the recruitment was considered, otherwise the first instance within thirty days of recruitment was used.

### Statistical analyses

Differences in demographic and clinical characteristics between the PDAC and PnC groups were assessed using Pearson’s Chi-square test or Kruskal-Wallis rank sum test, as appropriate. Predictor variables with data missing in more than 25% of the final study population (*N*=704) were excluded from further analyses. We conducted the preliminary observational association study and the subsequent development of the risk-prediction algorithms on 75% of the data (derivation set). We then assessed the prediction algorithms on the remaining 25% (validation set). Both derivation and validation sets had similar proportions of PDAC and PnC patients. Multiple imputation by chained equations method was used to replace missing values for predictor variables.^22,23^ Five imputations were carried out separately in derivation and validation datasets. In the imputed datasets, results from each blood test were stratified by the known normal range of respective pathology test guidelines (Supplementary Table 4) and included in the analyses as categorical variables with three possible categories: low, normal and high. To examine the PDAC risk associated with individual predictor variables, the effect size was evaluated with odds ratios (OR) with 95% confidence intervals (CI), using multivariable regression models with a binomial distribution. OR for individual predictor variables were obtained from independent regression models, controlled for potential confounding effect by demographic variables - age group, gender, and ethnicity. Rubin’s rules were used to combine the results across the imputed datasets.^22^

Several risk-prediction models were developed utilising the findings from observational association investigation. All predictor variables having at least one category with high statistical significance (p<0.01) after correction for multiple testing were retained to be used in the development of prediction models. Finally, with an aim to capture the effect of presenting symptoms and common blood tests in specific patient subgroups, we examined their interactions to encompass all demographic, comorbidity and lifestyle variables; all statistically significant two-way interactions (p<0.01) were subsequently included. Multivariate logistic regression models were fitted within the supervised machine learning setting. The outcome was the probability that a patient would develop PDAC, derived as a function of the predictor variables. Five variations of logistic regression (LR) models were fitted with: no penalty function, lasso regression with L1 regularisation penalty (LR-Lasso), ridge regression with L2 regularisation penalty utilising Bayesian Information Criterion (LR-Ridge), elastic net regression with L1/L2 regularisation penalty (LR-ElasticNet), and stepwise model selection by backward elimination utilising Akaike Information Criterion (LR-StepAIC). The average receiver operating characteristic (ROC) statistic from repeated 10-fold cross-validation training run was used as the metric to determine the optimal hyperparameters for the prediction models. The regression coefficients in the final LR, LR-Lasso, LR-Ridge and LR-ElasticNet models were pooled from preliminary models fitted to the imputed datasets. The final LR-StepAIC model was a regular LR model fitted with a reduced set of predictor variables, after applying a voting-based variable selection technique to the five LR-StepAIC models on the imputed datasets – variables were excluded if they did not appear in all the preliminary models.

Models’ performance were assessed based on the pooled outcome probability from five multiply imputed validation datasets. The best model was chosen based on the discrimination and calibration ability, which were measured by calculating the area under the receiver operating characteristic curve (AUROC) statistic^24^ and Spiegelhalter Z-test^25^ respectively. Once the best model was selected, it was applied on the full dataset to extract the *F*-scores and decide on the optimal probability threshold for identifying potential high-risk PDAC cases. *F*_*1*_ score is the harmonic mean of precision (positive predictive value) and recall (sensitivity) with equal importance. *F*_*2*_ score attaches twice as much importance to recall as precision, which is an important consideration for this study to distinguish PDAC patients from non-malignant pancreatic condition patients rather than “otherwise healthy” control population.

All statistical analyses and visualisations were performed in R (version 3.5.1).

## Results

### Descriptive analyses

Overall, the study included 344 patients with a primary diagnosis of PDAC and 360 patients with other pancreatic non-malignant conditions (PnC) (Figure 1). The PnC group consisted of patients diagnosed with chronic pancreatic conditions (CP) including chronic pancreatitis (n=113, 31%), benign tumour (n=86, 24%), and pancreatic cyst and pseudocyst (n=57, 16%); the rest of the patients (n=104, 29%) had acute pancreatitis or other benign diseases without progression to chronic conditions. Supplementary Table 5 presents baseline characteristics of the two groups. Both groups were male dominated, but the representation is notably higher in the PnC group (p=0.019). PDAC patients were significantly older (median 68 years, interquartile range (IQR) 60–75 years) compared to PnC (median 55, IQR 45-65.2, p<0.001), and had more prevalent comorbidities, particularly diabetes (p=0.002 and high blood pressure (p<0.001). Both groups had similar majority representation from White ethnic backgrounds, but South Asian representation was notably lower in the PDAC group (4.9%) than in the PnC group (10.3%) (p<0.001). At the time of recruitment (closer to the time of index diagnosis), PDAC patients had relatively lower BMI (median 24.2, IQR 22.1-27.5) compared to PnC patients (median 25.5, IQR 22.8-29.3, p=0.002). Histories of smoking and alcohol consumption were marginally less in PDAC group compared to PnC group, with notably lower current smokers (18.6% vs 27.2%) and past drinkers (12.8% vs 21.1%) among PDAC patients.

In the year prior to recruitment, jaundice and weight loss were reported more frequently by PDAC cases, while pain, nausea, vomiting and diarrhoea were more common among PnC patients (Supplementary Table 6). Although the incidence of pruritus was overall low among pancreatic patients, it was reported around four times more frequently by PDAC cases than PnC patients (p=0.005). The biochemical profiles of PDAC patients show significant association with higher levels of ALT, MCV, PLT, NEUT, CRP (all p<0.01) and lower levels of ALB, AMY, CREA, SOD, RBC, HB and LYMP (all p<0.01). PDAC patients demonstrated nearly two times higher levels of ALP (median 162.5 vs 86) and TBIL (median 14.5 vs 8.0) than PnC patients (Supplementary Table 6). Unsurprisingly, being a known pancreatic cancer biomarker, CA19-9 levels were very high among PDAC patients (median 294.0, IQR 63.8-1144.0) compared to the PnC group (median 15.9, IQR 9.0-35.0, p<0.001) (Supplementary Table 6).

### Observational association study

The derivation dataset was used to inspect the association between study variables and odds of PDAC in comparison to PnC (Figure 2). The dataset consisted of 258 PDAC patients and 270 PnC patients. The following predictor variables were excluded from the analyses due to missing data in more than 25% patients: AMY (77.6%), AST (62.4%), CA19-9 (46.6%), CA (40.1%), and CRP (31.4%). After controlling for demographic variables in the regression models, no comorbidity or lifestyle variables appeared to be independently associated with PDAC compared to PnC. Increased odds of PDAC was observed for increasing age group (OR between 3.6 and 10.1; p<0.001), jaundice and weight loss (nearly 3-fold; p<0.001). Compared to the normal levels of blood test results, increased odds of PDAC diagnosis were significantly associated with elevated levels of ALP (OR 2.9 [1.5-5.5]; p=0.003), ALT (OR 2.6 [1.5-4.4]; p=0.002), and TBIL (OR 4.8 [2.9-8.1]; p<0.001), and reduced levels of LYMP (OR 1.9 [1.2-3.0]; p=0.008), CREA (OR 3.1 [1.4-6.6]; p=0.008) and NA (OR 4.2 [1.6-10.6]; p=0.006). Unadjusted odds ratios are reported in Supplementary Figure 1. There was also evidence to suggest that PDAC risk associated with several predictors may vary with lifestyle or comorbidity status, particularly recent weight loss appearing to be a significant indicator of PDAC in hypertensive patients (OR 3.8 [1.6-8.9]; p=0.005) (Supplementary Table 7).

**Figure 2.**
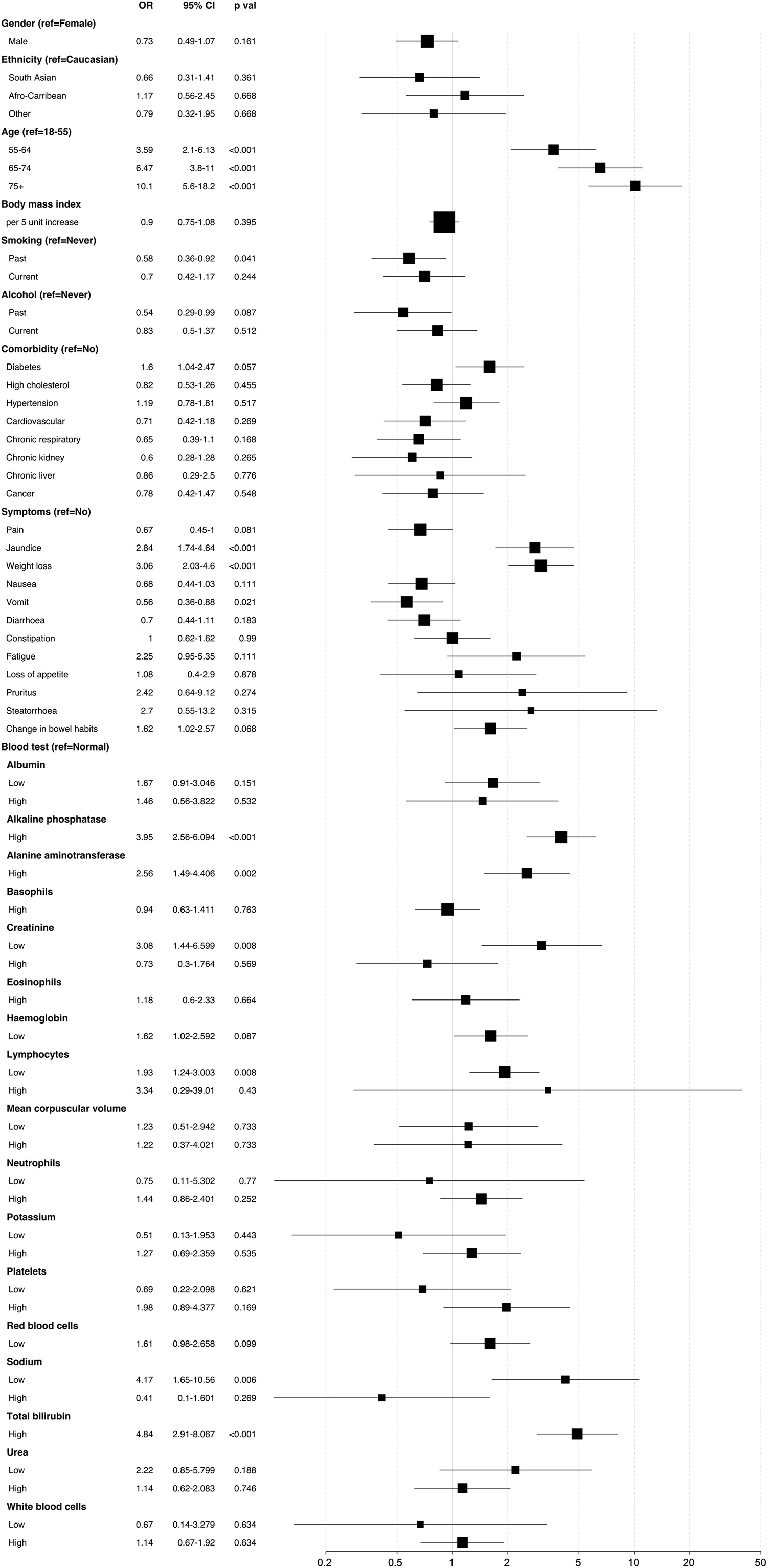
Forest plot showing association between study variables and odds of PDAC in comparison to non-malignant pancreatic disease group. The odds ratio (OR) and 95% confidence interval (CI) are derived from logistic regression model controlled for gender, ethnicity and age group. The reported p values are corrected for multiple testing via Benjamini-Hochberg method.

### Prediction models for differential diagnosis of PDAC

Statistically significant predictor variables and interactions from the observational association study were included to develop the prediction models under different algorithms: age group, jaundice, weight loss, ALP, ALT, CREA, LYMP, NA, TBIL, and hypertension – weight loss interaction.

The performances of the final models under different algorithms were checked in the multiply imputed validation dataset comprising of unique 86 PDAC cases and 90 PnC patients. The AUROC curves of all prediction models were identical, and significantly better than the null hypothesis area of 0.50, ranging between 0.88 and 0.89. (Table 1). However, the simplest LR model with no penalty function had the best calibration statistic (Spiegelhalter’s z=-1.25, p=0.21) (Table 1). Hence, LR model was selected as our final model (Table 2) indicating good differentiation of PDAC from the PnC cases whilst having close correspondence between predicted PDAC risk and observed outcome.

**Table 1.** Comparison of AUROC (area under the receiver operating characteristic) curve scores and Spiegelhalter’s z test statistics for all models on the validation dataset (n=176).

| Model | AUROC |  | Spiegelhalter's z test |  |
| --- | --- | --- | --- | --- |
|  | Area | 95% CI | Statistic | p value |
| LR | 0.88 | 0.83-0.93 | -1.25 | 0.21 |
| LR-StepAIC | 0.89 | 0.84-0.93 | -1.62 | 0.11 |
| LR-Ridge | 0.88 | 0.83-0.89 | -1.91 | 0.06 |
| LR-Lasso | 0.88 | 0.84-0.93 | -1.34 | 0.18 |
| LR-ElasticNet | 0.88 | 0.83-0.93 | -1.51 | 0.13 |

LR: logistic regression with no penalty function; LR-StepAIC: stepwise logistic regression with backward elimination utilising Akaike Information Criterion; LR-Ridge: ridge logistic regression with L2 regularisation penalty utilising Bayesian Information Criterion; LR-Lasso: lasso logistic regression with L1 regularisation penalty; LR-ElasticNet: elastic net logistic regression with L1/L2 regularisation penalty.

**Table 2.** Composition of the final PDAC risk prediction algorithm based on logistic regression with no penalty.

| | $\beta$<br>Coefficient | Standard<br>error (SE) | Odds ratio<br>(OR) | 95% confidence<br>interval (CI) |
| --- | --- | --- | --- | --- |
| **Intercept** | -2.27 | 0.26 | 0.1 | 0.06-0.17 |
| **Demographics** |  |  |  |  |
| *Age group (ref=<55)* |  |  |  |  |
| 55-64 | 0.89 | 0.27 | 2.43 | 1.42-4.15 |
| 65-74 | 1.52 | 0.28 | 4.59 | 2.68-7.87 |
| 75+ | 1.87 | 0.31 | 6.47 | 3.56-11.78 |
| **Symptoms (ref=No)** |  |  |  |  |
| Jaundiced | 0.67 | 0.25 | 1.96 | 1.2-3.21 |
| Weight loss | 0.33 | 0.26 | 1.39 | 0.84-2.3 |
| **Blood tests (ref=Normal)** |  |  |  |  |
| *Alkaline phosphatase* |  |  |  |  |
| Low | -0.5 | 0.64 | 0.61 | 0.17-2.14 |
| High | 0.76 | 0.24 | 2.14 | 1.34-3.4 |
| *Alanine aminotransferase* |  |  |  |  |
| High | -0.07 | 0.26 | 0.94 | 0.56-1.56 |
| *Creatinine* |  |  |  |  |
| Low | 0.75 | 0.36 | 2.12 | 1.04-4.31 |
| High | -0.6 | 0.42 | 0.55 | 0.24-1.26 |
| *Lymphocytes* |  |  |  |  |
| Low | 0.52 | 0.23 | 1.68 | 1.08-2.62 |
| High | 0.18 | 0.61 | 1.2 | 0.36-4 |
| *Sodium* |  |  |  |  |
| Low | 1.04 | 0.41 | 2.82 | 1.27-6.27 |
| High | -0.37 | 0.49 | 0.69 | 0.26-1.8 |
| *Bilirubin* |  |  |  |  |
| High | 0.88 | 0.28 | 2.42 | 1.39-4.21 |
| **Interactions** |  |  |  |  |
| *Hypertension* |  |  |  |  |
| without weight loss | -0.23 | 0.28 | 0.79 | 0.46-1.37 |
| with weight loss | 1.08 | 0.32 | 2.94 | 1.58-5.47 |

The maximum *F*_*2*_ score from the LR model applied to full dataset was obtained for probability threshold at 0.17, in which case 79.7% (n=561 of 704) of the study population would undergo urgent referral with a PDAC sensitivity of 95.6%. With a biomarker panel of 87.5% sensitivity^26^ applied, this could lead to early detection of around 83.7% (n=288 of 344) of PDAC patients (Figure 3). For a higher probability threshold of 0.40 from maximum *F*_*1*_ score, 54.1% (n=381) of the population would undergo urgent referral with early detection rate of 72%. Table 3 presents the sensitivity, specificity and positive predictive value for different probability cut-offs obtained from maximum *F*_*1*_ and *F*_*2*_ scores, and comparison with current National Institute for Health and Care Excellence (NICE) referral guidelines for suspected pancreatic cancer through specialist appointment within two weeks or urgent imaging within two weeks.^27^

**Figure 3.**
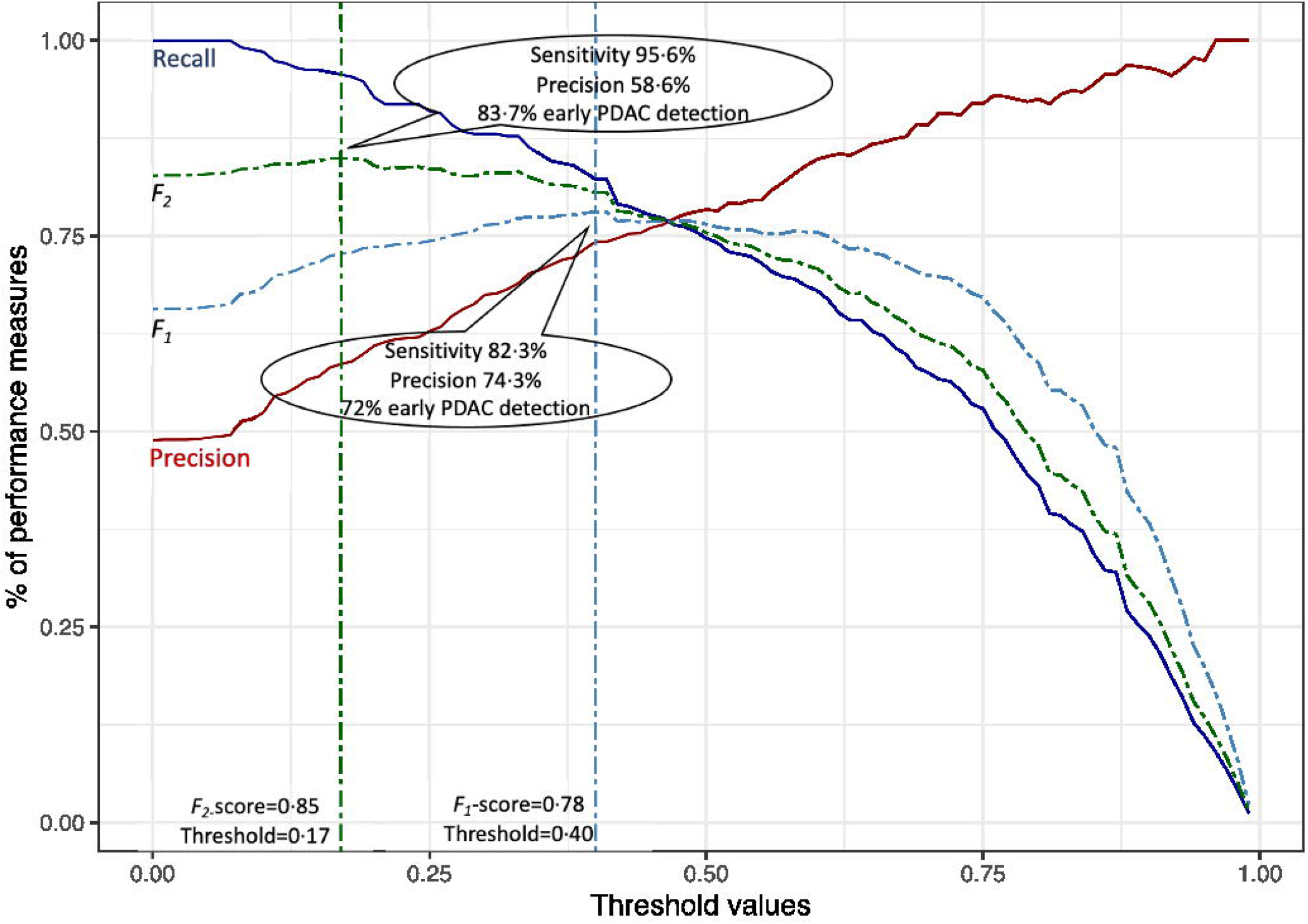
A performance measure graph with precision, recall, *F*_*1*_ and *F*_*2*_ of the logistic regression model with no penalty function with the resultant performance of the model at maximum *F*_*1*_ and *F*_*2*_ scores on the full dataset. The resultant percentage of early PDAC detection includes the use of a 87.5% sensitive biomarker.

**Table 3.** Performance measures of the logistic regression model with different evaluation criteria (maximum *F*_*1*_ and *F*_*2*_ score) and comparison with NICE referral guidelines for suspected pancreatic cancer (*N*=704).

|  | LR |  | Two week wait<sup>a</sup> | Urgent imaging<sup>b</sup> |
| --- | --- | --- | --- | --- |
| | $F_1$ criteria | $F_2$ criteria | | |
| Maximum $F$-score | 0.781 | 0.849 | 0.603 | 0.483-0.519 |
| Probability threshold | 0.400 | 0.170 | - | - |
| Sensitivity | 0.823 | 0.956 | 0.488 | 0.352-0.390 |
| Specificity | 0.728 | 0.356 | 0.875 | 0.894-0.900 |
| Precision | 0.743 | 0.586 | 0.789 | 0.771-0.779 |
| Proportion identified for referral | 0.541 | 0.797 | 0.274 | 0.223-0.244 |
| Maximum proportion of early detection of cancer<sup>c</sup> | 0.720 | 0.837 | 0.427 | 0.308-0.341 |

<sup>a</sup> According to NICE guideline for specialist appointment within two weeks for patients aged 40 or over with jaundice
<sup>b</sup> According to NICE guideline for urgent direct access CT scan (to be performed within 2 weeks), or an urgent ultrasound scan if CT is not available, for patients aged 60 or over with weight loss and any of the following: diarrhoea, pain, nausea, vomiting, constipation, new-onset diabetes. As distinction cannot be made between diabetes and new-onset diabetes from our data, results are represented as range derived from two sets of calculation: i) excluding new-onset diabetes from the formula; ii) diabetes variable as placeholder for new-onset diabetes.
<sup>c</sup> Considering a biomarker panel of 87.5% sensitivity

## Discussion

We conducted a case-control study on a patient cohort prospectively recruited at a pancreatic cancer-focused biobank. We identified features in pancreatic patients’ recent medical history that could separate putative pancreatic ductal adenocarcinoma cases from benign pancreatic conditions, which present similarly clinically and are included as differential diagnoses. We utilised the findings to develop and validate a new prediction algorithm for differential diagnosis. To our knowledge, this is the first prospectively recruited study holistically exploring the presenting symptoms and common laboratory test results of pancreatic disease patients, at the time point of their consultation at secondary/tertiary care, within the context of their demographic, lifestyle and comorbidity characteristics. The development dataset is well proportioned with a large number of PDAC patients comparable to other prospective studies.^14,28^ The final algorithm was shown to have good discrimination and calibration on a separate validation dataset.

The study reaffirms previously known associations with PDAC such as age, symptoms of jaundice and weight loss^9,18^ as well as raised serum bilirubin, deranged liver function tests, raised serum CA19-9 and raised inflammatory markers.^7,19^ PDAC is known to clinically present with obstructive jaundice due to either invasion or mass effect on the biliary tree causing deranged liver function tests (ALP, ALT and TBIL). Although other pancreatic cancers or benign pancreatic disease may present this way, PDAC is less prevalent than benign disease with a significantly worse mortality and morbidity. RBC count and Hb levels have been previously shown to be important when separating between PDAC, CP and healthy controls.^29^ While our study found both Hb and RBC to be independently associated with PDAC, both were attenuated to statistical non-significance after correction for demographic parameters. Further studies into the biology of low haemoglobin and RBC count in subgroups of PDAC patients are therefore warranted.

We have also found novel associations with PDAC when compared to other pancreatic disease which will likely translate to amplified associations when compared to healthy patients. Low sodium, which has previously not been reported in the literature for PDAC, may reflect undiagnosed SIADH (syndrome of inappropriate antidiuretic hormone) in PDAC patients or perhaps may be due to pathophysiology of the cancer.^30^ This will need further investigation into clinical correlation and may allow an avenue into understanding the disease process better. Raised ALP, found to be significantly associated with PDAC, may reflect biliary tree injury secondary to biliary obstruction in PDAC patients. It is not classically a blood test used to differentiate between PDAC and benign disease.

Unlike what the literature suggests, we found no association between higher BMI and PDAC.^6,14,18^ This could be due to the use of benign pancreatic disease patients as comparison cohort, suggesting that obesity is likely to be associated with pancreatic conditions in general rather than severity of the disease.^31^ We used patients’ most recent pre-diagnostic BMI rather than historical BMI trend, which could have also masked PDAC patients’ transition from higher to lower BMI status. A further novel finding was the association of low creatinine with PDAC, this may be due to damage to the liver through obstructive jaundice or disease associated sarcopenia and cachexia.

Our final prediction algorithm shows that age (over 55), weight loss in hypertensive patients, recent symptoms of jaundice, high serum bilirubin, high serum ALP, low serum creatinine, low lymphocyte counts and low serum sodium are the most important features when identifying putative PDAC cases from less severe pancreatic conditions. The power of machine learning techniques allowed us to identify multiple weak indicators, which became only predictive for differential diagnosis when used in complex combination with each other. We have shown that using the separation probability threshold of 0.17 derived from maximum *F*_*2*_-score could lead to early detection of around 84% of the PDAC cases within our study cohort. This would optimise the use of the urgent referral pathway and/or costly investigative procedures through maximising identification of potential PDAC cases at the cost of slightly increased false positive detection of non-cancerous pancreatic patients. With the separation probability threshold set at 0.40, our algorithm has sensitivity of 82% and *F*_*1*_-score of 78%. This is still better than known diagnostic biomarkers (median sensitivity 79%).^32^ It is important to note that prediction algorithms are often developed to identify PDAC or pancreatic cancer in general population with significant representation of healthy controls,^6,14,18,29,33^ whereas our comparison cohort consisted of benign pancreatic disease patients who demonstrate similar clinical manifestations as the pancreatic cancer patients. Hence, a direct performance comparison with our model may not be appropriate, yet it is comparable to regression based established prediction algorithms such as QCancer Pancreas (AUROC: 84-92% vs 85.6%)^33^ or even ensemble-learning based complex algorithm (AUROC: 96%; *F*_*1*_-score: 64% vs 78%).^29^ The established protocols such as current NICE guideline in the UK for urgent referral to cancer clinics or urgent imaging-based investigation within two weeks^27^ shows a very low sensitivity between 35% and 49% had these been applied to our study population of pancreatic patients, and that possibly explains why a vast majority of pancreatic cancer cases are still diagnosed as a result of an emergency presentation to hospital.^18^

Risk prediction algorithms have been an important tool in both primary, secondary or tertiary care and have helped patients with these time sensitive diseases to be diagnosed and treated earlier, improving survival and quality of life.^34^ Our prediction algorithm may become a useful adjunct to the clinical skills of primary care physicians as it includes widely available and easy to request blood tests, symptoms that the patients should be able to recognise when they occur, well-recorded medical comorbidities, and demographic details of patients. We hope that its use may help appropriately triage patients, who may not present with jaundice or obviously deranged liver function tests, towards a two-week wait clinic appointment at a specialised tertiary centre for expedited diagnosis and treatment. This has also created the ground-work to develop an easy to understand, utilisable risk score since it was trained through logistic regression rather than neural networks or ensemble learning models. However, we cannot ignore the need for a further rigorous external validation before clinical implementation.

Key strengths of our study include the study design, with adherence to the STROBE guidelines and TRIPOD statement,^35,36^ a prospectively recruited cohort as well as high quality clinically relevant data which was recorded through participant interviews and EHR data extraction followed by independent verification by trained clinicians. This minimises recall, response and information bias. Another strength is the healthy representation of patients from non-White origin (21%), particularly South Asian and Afro-Caribbean, thereby increasing the generalisability of the results. This is the only study in our knowledge that has compared PDAC with other pancreatic disease at the time of presentation since these two groups of diseases are common differential diagnoses with very similar clinical and laboratory profiles. As the recruited cohort of patients have consented for tissue and blood samples to be given to the biobank, further biological studies can be performed based on our analysis with future improvement of the prediction model to include novel biomarkers, tissue histology, imaging results, genomic profiling, survival and operative findings.

One key limitation of the study is amount of missing data for blood test variables. We used a broader time window to collect participants’ blood test results; yet not all patients who visited the specialist HPB centre with suspected pancreatic conditions had those blood tests performed within the window. We attempted to minimise the impact by first removing variables with missing data in >25% patients and then conducting analyses on multiple imputed data. The prediction algorithm’s performance may improve further with completeness of laboratory test variables such as CA19-9, carcinoembryonic antigen and amylase. We also used the reported blood test results as reported with no reference or adjustment to any ongoing medical interventions that could have affected the results; although most PDAC patients were recruited before diagnosis and intervention. Another limitation of the study is the risk of unmeasured residual confounding. For example, the confounding effect of diabetes on PDAC risk could be different if participants with new-onset diabetes could be separated from those with long-standing diabetes. The overall study population was smaller in comparison to other retrospective cohort studies focusing on predicting pancreatic cancer risk in the general population. However, considering the study objective of separating a low-prevalence difficult-to-diagnose malignant disease from its non-malignant counterpart, our study had a healthy and balanced representation of cases and controls with reliable and accurate data. In order to gain sufficient power to take full advantage of machine learning, we acknowledge the need of a much larger prospective study. This could be logistically challenging to achieve. However, biobanks such as the Barts Pancreas Tissue Bank can support towards the goal with continuous recruitment of patients following a standardised and ethically approved protocol, and also particularly useful in providing granularity of data that can address important questions in cancer research.

In summary, this study has shown novel clinical associations with PDAC as well as generating a simple and accurate prediction algorithm to guide primary care and secondary care physicians when referring suspected pancreatic patients to specialist HPB services. There is strong evidence that current UK guidelines for urgent referral are underperforming for this patient group and integration of the common laboratory tests highlighted here will improve the accuracy of such guidance. Further research is warranted, utilising both primary and secondary care data of patients to get a full picture of their medical journey, to identify more clinically significant parameters for the prediction algorithm supporting the identification of PDAC patients in the most timely and appropriate way.

## Data Availability

All data produced in the present study are available upon reasonable request to Barts Pancreas Tissue Bank.

## Abbreviations

AUROC: (area under the receiver operating characteristic)
BHNT: (Barts Health NHS Trust)
BMI: (body mass index)
BPTB: (Barts Pancreas Tissue Bank)
CDST: (cancer decision support tool)
CI: (confidence interval)
EHR: (electronic health records)
HPB: (hepato-pancreatico-biliary)
LR: (logistic regression)
NICE: (National Institute for Health and Care Excellence)
OR: (odds ratio)
PDAC: (pancreatic ductal adenocarcinoma)
PnC: (non-malignant pancreatic diseases)
ROC: (receiver operating characteristic)

## Acknowledgements

We thank PCRFTB Operations Group (Maggie Blanks, Brian Davidson, Mo Abu’Hilal, Ali Arshad, Zahir Soonawalla, Deep Malde, Bilal Al-Sarireh, Satyajit Bhattacharya, Stuart Robinson) and surgeons and oncologist at Barts Health NHS Trust (Robert Hutchins, Ajit Abraham, Vincent Yip, Deepak Hariharan, David Propper, Sarah Slater), BPTB Tissue Access Committee (chaired by Richard Grose) and PCRFTB Tissue Access Committee (chaired by Ian Hart) for their support in facilitating this study.

We acknowledge the contribution to the research made by several members of the BPTB and PCRFTB teams (Archana Ambily, Thomas Dowe, Opeoluwa Banwo, Jacek Marzec, Stefano Pirro, Mingshing Yang, Rory Smith, Shreya Sharma, Michael Ebose, Catherine Graham) and clinical research fellows (Andrew Ang, Athena Michaelides, Shanthini Crusz) at Barts Cancer Institute through insightful medical and scientific discussion.

Finally, we thank all patients for donating the samples and providing the clinical information to the Tissue Bank.

