## Supplemental data for "Development of a Model for Differentiating PDAC from Benign Pancreatic Conditions: A Prospective Case-control Study"

**Supplementary data**

**Contents**

**Table 1** A list of diagnoses that fall under the three diagnostic groups with their associated ICD-10 and ICD-0-3 codes. 2

**Table 2** Frequency of diagnoses in the study patient groups. 3

**Table 3** Clinical variables considered for the observational association study. Statistically significant variables were then used in the development of the risk prediction models. 4

**Table 4** Reference ranges of the blood tests used to determine the high and low ranges for the corresponding tests. 5

**Table 5** Baseline characteristics of the study groups. 6

**Table 6** Observed pre-diagnostic symptoms and common blood test results of the study groups. 8

**Table 7** Modified odds of PDAC associated with different predictor variables among different participant subgroups. Only statistically significant interactions. 11

**Figure 1** Forest plot showing crude association between study variables and odds of PDAC in comparison to non-malignant pancreatic disease group. 12

**Supplementary table 1.** A list of diagnoses that fall under the three diagnostic groups with their associated ICD-10 and ICD-0-3 codes.

| ICD-10 | ICD-0-3 | Diagnosis |
|---|---|---|
| **Pancreatic ductal adenocarcinoma (PDAC)** | | |
| C25 | 8500/3 | Infiltrating duct carcinoma NOS |
| **Pancreas cancers (PC)** | | |
| C25 | 8550/3 | Acinar cell carcinoma |
| C25 | 8551/3 | Acinar cell cystadenocarcinoma |
| C25 | 8453/3 | Intraductal papillary-mucinous carcinoma, invasive |
| C25 | 8470/3 | Mucinous cystadenocarcinoma NOS |
| C25 | 8971/3 | Pancreatoblastoma |
| C25 | 8441/3 | Serous cystadenocarcinoma, NOS |
| C25 | 8452/3 | Solid pseudopapillary carcinoma |
| C25 | 8240/3 | Carcinoid tumor, NOS |
| C25 | 8240/3 | Typical carcinoid |
| C25 | 8240/3 | Bronchial adenoma, carcinoid |
| C25 | 8249/3 | Atypical carcinoid tumor |
| C25 | 8246/3 | Neuroendocrine carcinoma, NOS |
| C25 | 8241/3 | Enterochromaffin cell carcinoid |
| C25 | 8153/3 | Gastrinoma. malignant |
| C25 | 8152/3 | Glucagonoma. malignant |
| C25 | 8151/3 | Insulinoma. malignant |
| C25 | 8156/3 | Somatostatinoma malignant |
| C25 | 8155/3 | Vipoma malignant |
| C25 | 9260/3 | Ewing sarcoma |
| C25 | 8806/3 | Desmoplastic small round cell tumor |
| C25 | 8453/2 | Intraductal papillary-mucinous carcinoma, non-invasive |
| C25 | 8470/2 | Mucinous cystadenocarcinoma, non-invasive |
| **Non-malignant pancreatic conditions** | | **(PnC)** |
| K85.2 | | Alcohol-induced acute pancreatitis |
| K85.3 | | Drug-induced acute pancreatitis |
| K85.0 | | Idiopathic acute pancreatitis |
| K85.9 | | Acute pancreatitis, unspecified |
| K85.1 | | Biliary acute pancreatitis |
| K86.2 | | Cyst of pancreas |
| K86.3 | | Pseudocyst of pancreas |
| K83.4 | | Spasm of sphincter of Oddi |
| Q45.3 | | Other congenital malformations of pancreas and pancreatic duct |
| C25 | 8150/0 | Pancreatic endocrine tumor, benign |
| C25 | 8150/0 | Islet cell adenomatosis |
| C25 | 8453/0 | Intraductal papillary-mucinous adenoma |
| C25 | 8470/0 | Mucinous cystadenoma, NOS |
| | 8441/0 | Serous cystadenoma, NOS |
| | 8441/0 | Serous microcystic adenoma |
| | 9170/0 | Lymphangioma, NOS |
| | 8850/0 | Lipoma, NOS |

**Supplementary table 2.** Frequency of diagnoses in the study patient groups.

| Diagnosis | Count (%) |
|---|---|
| **PDAC** | **344(45·1%)** |
| Pancreatic ductal adenocarcinoma | 344 |
| **Non-malignant pancreatic conditions (PnC)** | **360(47·2%)** |
| Chronic pancreatitis | 113 |
| Cyst and pseudocyst of pancreas | 57 |
| Intraductal papillary mucinous adenoma | 36 |
| Acute pancreatitis - unspecified | 32 |
| Biliary acute pancreatitis | 30 |
| Serous cystadenoma | 22 |
| Alcohol/drug-induced acute pancreatitis | 19 |
| Pancreatic endocrine tumor - benign | 14 |
| Mucinous cystadenoma | 11 |
| Spasm of sphincter of oddi | 9 |
| Idiopathic acute pancreatitis | 9 |
| Congenital malformation of pancreas and pancreatic duct | 4 |
| Serous microcystic adenoma | 3 |
| Autoimmune pancreatitis | 1 |
| **Pancreas cancer** | **58(7·6%)** |
| Neuroendocrine Carcinoma | 20 |
| Intraductal papillary-mucinous carcinoma - non-invasive | 11 |
| Insulinoma | 9 |
| Solid pseudopapillary carcinoma | 6 |
| Intraductal papillary mucinous carcinoma | 5 |
| Mucinous cystadenocarcinoma - non-invasive | 2 |
| Glucagonoma | 2 |
| Acinar cell carcinoma | 1 |
| Carcinoid tumour | 1 |
| Serous cystadenocarcinoma | 1 |
| **Total** | **762(100%)** |

**Supplementary table 3.** Clinical variables considered for the observational association study. Statistically significant variables were then used in the development of the risk prediction models.

| Demographics | Symptoms | Medical history | Lifestyle history | Laboratory results |
|---|---|---|---|---|
| Gender | Pain | Diabetes | Smoking | Urea (UREA) |
| Age | Jaundice | Cholesterol | Alcohol | Potassium (K) |
| Ethnicity | Weight loss | Hypertension | Body mass index (BMI) | Sodium (NA) |
| | Nausea | Cardiovascular | | Total calcium (CA) |
| | Vomiting | Chronic respiratory | | Creatinine (CREAT) |
| | Diarrhoea | Chronic kidney | | Total bilirubin (TBIL) |
| | Constipation | Chronic liver | | Alkaline phosphatase (ALP) |
| | Fatigue | Cancer | | Alanine aminotransferase (ALT) |
| | Loss of appetite | | | Aspartate transaminase (AST) |
| | Pruritus | | | Albumin (ALB) |
| | Steatorrhoea | | | Amylase (AMY) |
| | Change in bowel habits | | | Carbohydrate antigen (CA19-9) |
| | | | | Haemoglobin (HB) |
| | | | | Red blood cell count (RBC) |
| | | | | Mean corpuscular volume (MCV) |
| | | | | Platelets (PLT) |
| | | | | White blood cell count (WBC) |
| | | | | Basophils (BASO) |
| | | | | Eosinophils (EOSI) |
| | | | | Lymphocytes (LYMP) |
| | | | | Neutrophils (NEUT) |
| | | | | C-reactive protein (CRP) |

**Supplementary table 4.** Reference ranges of the blood tests used to determine the high and low ranges for the corresponding tests.

| **Blood test** | **Reference range** |
|---|---|
| Haemoglobin (HB) | 1.0 - 180 g/L (male)<br>1.5 - 165 g/L (female) |
| White Blood Cell Count (WBC) | 4.0 - 11.0 $10^{9}$ /L |
| Red Blood Cell Count (RBC) | 4.5 - 6.5 $10^{12}$ /L (male)<br>3.8 - 5.8 $10^{12}$ /L (female) |
| Mean Corpuscular Volume (MCV) | 0 - 100 fL |
| Platelet count (PLT) | 150 - 400 $10^{9}$ /L |
| Neutrophils (NEUT) | 2 - 7.5 $10^{9}$ /L |
| Lymphocytes (LYMP) | 1.5 - 4.5 $10^{9}$ /L |
| Eosinophils (EOSI) | 0 - 0.4 $10^{9}$ /L |
| Basophils (BASO) | 0 - 0.1 $10^{9}$ /L |
| Total bilirubin (TBIL) | 1 - 17 µmol/L |
| Alkaline phosphatase (ALP) | 30 - 130 U/L |
| Aspartate transaminase (AST) | 10 - 40 U/L |
| Alanine transaminase (ALT) | 1 - 50 U/L |
| Amylase (AMY) | 0 - 118 U/L |
| Albumin (ALB) | 35 - 50 g/L |
| CA 19-9 (C199) | 0 - 37 kU/L |
| C-reactive protein (CRP) | 0 - 5 mg/L |
| Creatinine (CREAT) | 0 - 120 µmol/L |
| Urea (UREA) | 2.5 - 7 mmol/L |
| Sodium (NA) | 133 - 146 mmol/L |
| Potassium (K) | 3.5 - 4.5 mmol/L |
| Total calcium (CA) | 2.2 - 2.6 mmol/L |

**Supplementary table 5:** Baseline characteristics of the study groups.

| | PDAC (n=344) | PnC (n=360) | Total (N=704) | p value[a] |
|---|---|---|---|---|
| **Gender** | | | | 0.019 |
| Female | 162 (47.1%) | 138 (38.3%) | 300 (42.6%) | |
| Male | 182 (52.9%) | 222 (61.7%) | 404 (57.4%) | |
| **Ethnicity** | | | | < 0.001 |
| Caucasian | 272 (79.1%) | 241 (66.9%) | 513 (72.9%) | |
| South Asian | 17 (4.9%) | 37 (10.3%) | 54 (7.7%) | |
| Afro-Caribbean | 30 (8.7%) | 27 (7.5%) | 57 (8.1%) | |
| Other | 15 (4.4%) | 20 (5.6%) | 35 (5.0%) | |
| Unknown | 10 (2.9%) | 35 (9.7%) | 45 (6.4%) | |
| **Age** | | | | < 0.001 [b] |
| Median (IQR) | 68.0 (60.0-75.0) | 55.0 (45.0-65.2) | 62.0 (51.0-72.0) | |
| **Body mass index (BMI)** | | | | 0.002 [b] |
| Median (IQR) | 24.2 (22.1-27.5) | 25.5 (22.8-29.3) | 25.0 (22.3-28.2) | |
| Unknown | 71 (20.6%) | 85 (23.2%) | 156 (22.2%) | |
| **Smoking** | | | | 0.022 |
| Never | 160 (46.5%) | 136 (37.8%) | 296 (42.0%) | |
| Past | 95 (27.6%) | 94 (26.1%) | 189 (26.8%) | |
| Current | 64 (18.6%) | 98 (27.2%) | 162 (23.0%) | |
| Unknown | 25 (7.3%) | 32 (8.9%) | 57 (8.1%) | |
| **Drinking** | | | | 0.018 |
| Never | 129 (37.5%) | 109 (30.3%) | 238 (33.8%) | |
| Past | 44 (12.8%) | 76 (21.1%) | 120 (17.0%) | |
| Current | 140 (40.7%) | 140 (38.9%) | 280 (39.8%) | |
| Unknown | 31 (9.0%) | 35 (9.7%) | 66 (9.4%) | |
| **Diabetes** | | | | 0.002 |
| Yes | 120 (34.9%) | 87 (24.2%) | 207 (29.4%) | |
| **Cholesterol** | | | | 0.073 |
| Yes | 113 (32.8%) | 96 (26.7%) | 209 (29.7%) | |
| **Hypertension** | | | | <0.001 |
| Yes | 158 (45.9%) | 102 (28.3%) | 260 (36.9%) | |
| **Cardiovascular disease** | | | | 0.054 |
| Yes | 70 (20.3%) | 51 (14.2%) | 121 (17.2%) | |
| Unknown | 1 (0.3%) | 0 (0.0%) | 1 (0.1%) | |
| **Chronic respiratory disease** | | | | 0.592 |
| Yes | 54 (15.7%) | 57 (15.8%) | 111 (15.8%) | |
| Unknown | 1 (0.3%) | 0 (0.0%) | 1 (0.1%) | |
| **Chronic kidney disease** | | | | 0.292 |
| Yes | 19 (5.5%) | 28 (7.8%) | 47 (6.7%) | |
| Unknown | 1 (0.3%) | 0 (0.0%) | 1 (0.1%) | |
| **Chronic liver disease** | | | | 0.526 |

| | | | | |
|---|---|---|---|---|
| Yes | 11 (3.2%) | 14 (3.9%) | 25 (3.6%) | |
| Unknown | 1 (0.3%) | 0 (0.0%) | 1 (0.1%) | |
| **Past history of Cancer** | | | | 0.017 |
| Yes | 42 (12.2%) | 25 (6.9%) | 67 (9.5%) | |

[a] Differences between groups evaluated by the $\chi 2$ test, unless otherwise stated.

[b] Differences between groups evaluated by Kruskal-Wallis rank sum test.

PDAC, pancreatic ductal adenocarcinoma; PnC: non-malignant pancreatic diseases; IQR, interquartile range

**Supplementary table 6.** Observed pre-diagnostic symptoms and common blood test results of the study groups

| | PDAC (n=344) | PnC (n=360) | Total (N=704) | p value |
|---|---|---|---|---|
| **Symptoms[a]** | | | | |
| **Pain** | | | | <0.001 |
| Yes | 175 (50.9%) | 262 (72.8%) | 437 (62.1%) | |
| **Jaundiced** | | | | <0.001 |
| Yes | 169 (49.1%) | 57 (15.8%) | 226 (32.1%) | |
| **Weight loss** | | | | <0.001 |
| Yes | 204 (59.3%) | 140 (38.9%) | 344 (48.9%) | |
| **Nausea** | | | | 0.009 |
| Yes | 95 (27.6%) | 136 (37.8%) | 231 (32.8%) | |
| Unknown | 0 (0.0%) | 1 (0.3%) | 1 (0.1%) | |
| **Vomiting** | | | | <0.001 |
| Yes | 62 (18.0%) | 124 (34.4%) | 186 (26.4%) | |
| Unknown | 0 (0.0%) | 1 (0.3%) | 1 (0.1%) | |
| **Diarrhoea** | | | | 0.021 |
| Yes | 61 (17.7%) | 93 (25.8%) | 154 (21.9%) | |
| Unknown | 1 (0.3%) | 0 (0.0%) | 1 (0.1%) | |
| **Constipation** | | | | 0.712 |
| Yes | 64 (18.8%) | 78 (21.2%) | 142 (20.1%) | |
| Unknown | 4 (1.2%) | 4 (1.1%) | 8 (1.1%) | |
| **Fatigue** | | | | 0.076 |
| Yes | 25 (7.3%) | 15 (4.2%) | 40 (5.7%) | |
| **Loss of appetite** | | | | 0.910 |
| Yes | 12 (3.5%) | 12 (3.3%) | 24 (3.4%) | |
| **Pruritus** | | | | 0.005 |
| Yes | 16 (4.7%) | 4 (1.1%) | 20 (2.8%) | |
| **Steatorrhea** | | | | 0.208 |
| Yes | 9 (2.6%) | 4 (1.1%) | 13 (1.8%) | |
| Unknown | 0 (0.0%) | 1 (0.3%) | 1 (0.1%) | |
| **Change in bowel habits** | | | | 0.077 |
| Yes | 88 (25.6%) | 72 (20.0%) | 160 (22.7%) | |
| **Blood tests[b]** | | | | |
| **Haemoglobin (HB)** | | | | <0.001 |
| Median (IQR) | 120.5 (100.0-132.5) | 130.0 (117.0-142.0) | 125.0 (106.5-39.9) | |
| Unknown | 21 (6.1%) | 53 (14.7%) | 74 (10.5%) | |
| **Red blood cell count (RBC)** | | | | <0.001 |
| Median (IQR) | 4.1 (3.7-4.6) | 4.5 (4.2-4.9) | 4.3 (3.9-4.8) | |
| Unknown | 51 (14.8%) | 100 (27.8%) | 151 (21.4%) | |

| | | | | |
|---|---|---|---|---|
| **Mean corpuscular volume (MCV)** | | | | <0.001 |
| Median (IQR) | 89.4 (85.5-93.9) | 87.8 (84.3-91.1) | 88.9 (84.8-92.6) | |
| Unknown | 51 (14.8%) | 100 (27.8%) | 151 (21.4%) | |
| **Platelets (PLT)** | | | | <0.001 |
| Median (IQR) | 287.0 (227.0-376.5) | 261.5 (214.8-311.2) | 273.0 (219.0-351.0) | |
| Unknown | 21 (6.1%) | 56 (15.6%) | 77 (10.9%) | |
| **White blood cell count (WBC)** | | | | 0.054 |
| Median (IQR) | 8.4 (6.8-10.4) | 7.9 (6.4-9.9) | 8.1 (6.7-10.2) | |
| Unknown | 21 (6.1%) | 56 (15.6%) | 77 (10.9%) | |
| **Neutrophils (NEUT)** | | | | 0.001 |
| Median (IQR) | 5.4 (4.3-7.4) | 4.9 (3.6-6.6) | 5.1 (3.9-7.0) | |
| Unknown | 51 (14.8%) | 100 (27.8%) | 151 (21.4%) | |
| **Lymphocytes (LYMP)** | | | | <0.001 |
| Median (IQR) | 1.7 (1.2-2.3) | 1.9 (1.5-2.4) | 1.8 (1.4-2.4) | |
| Unknown | 51 (14.8%) | 100 (27.8%) | 151 (21.4%) | |
| **Basophils (BASO)** | | | | 0.060 |
| Median (IQR) | 0 (0.0- 0.1) | 0 (0.0-0.1) | 0 (0.0-0.1) | |
| Unknown | 51 (14.8%) | 100 (27.8%) | 151 (21.4%) | |
| **Eosinophils (EOSI)** | | | | 0.467 |
| Median (IQR) | 0.1 (0.1-0.2) | 0.1 (0.1-0.3) | 0.1 (0.1-0.2) | |
| Unknown | 51 (14.8%) | 100 (27.8%) | 151 (21.4%) | |
| **C-reactive protein (CRP)** | | | | < 0.001 |
| Median (IQR) | 20.0 (7.0-55.0) | 7.0 (4.0, 38.0) | 14.0 (5.0-48.8) | |
| Unknown | 80 (23.3%) | 141 (39.2%) | 221 (31.4%) | |
| **Total bilirubin (TBIL)** | | | | <0.001 |
| Median (IQR) | 14.5 (6.0, 47.5) | 8 (5.8, 12.0) | 10 (6.0, 20.0) | |
| Unknown | 21 (6.1%) | 61 (16.9%) | 82 (11.6%) | |
| **Alkaline phosphatase (ALP)** | | | | <0.001 |
| Median (IQR) | 162.5 (93.5, 312.8) | 86.0 (69.0, 117.0) | 107.0 (77.8, 212.0) | |
| Unknown | 20 (5.8%) | 60 (16.7%) | 80 (11.4%) | |
| **Aspartate aminotransferase (AST)** | | | | 0.041 |
| Median (IQR) | 53.0 (30.0-96.2) | 41.0 (23.0-73.0) | 47.0 (26.0-86.0) | |
| Unknown | 168 (48.8%) | 271 (75.3%) | 439 (62.4%) | |
| **Alanine transaminase (ALT)** | | | | <0.001 |
| Median (IQR) | 34.2 (19.0-8.0) | 21.0 (15.0-41.8) | 27.0 (17.0-58.0) | |
| Unknown | 20 (5.8%) | 58 (16.1%) | 78 (11.1%) | |
| **Albumin (ALB)** | | | | <0.001 |
| Median (IQR) | 42 (38.0-45.0) | 44.0 (41.0-47.0) | 43 (39.0-46.0) | |
| Unknown | 20 (5.8%) | 56 (15.6%) | 76 (10.8%) | |

| | | | | |
|---|---|---|---|---|
| **Amylase (AMY)** | | | | <0.001 |
| Median (IQR) | 43.0 (25.2-76.0) | 90.0 (57.5-153.2) | 61.0 (32.0-110.0) | |
| Unknown | 254 (73.8%) | 292 (81.1%) | 546 (77.6%) | |
| **CA19-9** | | | | <0.001 |
| Median (IQR) | 294.0 (63.8-1144.0) | 15.9 (9.0-35.0) | 115.2 (19.3-670.5) | |
| Unknown | 81 (23.5%) | 247 (68.6%) | 328 (46.6%) | |
| **Total calcium (CA)** | | | | 0.089 |
| Median (IQR) | 2.3 (2.2-2.4) | 2.2 (2.2-2.4) | 2.3 (2.2-2.4) | |
| Unknown | 96 (27.9%) | 186 (51.7%) | 282 (40.1%) | |
| **Sodium (NA)** | | | | <0.001 |
| Median (IQR) | 138.5 (135.9-141.0) | 141.0 (139.0-142.0) | 140.0 (137.0-142.0) | |
| Unknown | 24 (7.0%) | 54 (15.0%) | 78 (11.1%) | |
| **Potassium (K)** | | | | 0.018 |
| Median (IQR) | 4.4 (4.1-4.8) | 4.3 (4.1-4.6) | 4.4 (4.1-4.7) | |
| Unknown | 24 (7.0%) | 52 (14.4%) | 76 (10.8%) | |
| **Creatinine (CREA)** | | | | 0.006 |
| Median (IQR) | 69 (56.0-84.0) | 74.0 (62.0-87.0) | 71 (58.5-86.0) | |
| Unknown | 24 (7.0%) | 55 (15.3%) | 79 (11.2%) | |
| **Urea (UREA)** | | | | 0.080 |
| Median (IQR) | 5 (3.9-6.3) | 4.8 (3.7-6.0) | 4.9 (3.7-6.1) | |
| Unknown | 24 (7.0%) | 60 (16.7%) | 84 (11.9%) | |

[a] Differences between groups evaluated by the $\chi 2$ test.

[b] Differences between groups evaluated by Kruskal-Wallis rank sum test.

PDAC, pancreatic ductal adenocarcinoma; PnC: non-malignant pancreatic diseases; IQR, interquartile range

**Supplementary table 7.** Modified odds of PDAC associated with different predictor variables among different participant subgroups. Only statistically significant interactions.

| **Modifier** | **Predictor** | | **p-inter** |
|---|---|---|---|
| | **OR [95% CI]** | **p-val** | |
| *Diabetes (ref=No)* | *Alanine transaminase (ref=Normal)* | | 0·010 |
| | High | | |
| Yes | 0·27 [0·08-0·86] | 0·04 | |
| *Smoking (ref=Never)* | *Chronic liver (ref=No)* | | 0·007 |
| | Yes | | |
| Past | 2·73 [0·14-54·1] | 0·711 | |
| Current | 1·86E-7 [0-Inf] | 0·977 | |
| *Hypertension (ref=No)* | *Weight loss (ref=No)* | | 0·006 |
| | Yes | | |
| Yes | **3·82 [1·63-8·93]** | **0·005** | |
| *Cancer (ref=No)* | *Vomit (ref=No)* | | 0·009 |
| | Yes | | |
| Yes | 9·29E6[0-Inf] | 0·98 | |

OR adjusted for gender, ethnicity and age group. The reported p values are corrected for multiple testing via Benjamini-Hochberg method.

Interaction between groups evaluated by the likelihood ratio test. Only statistically significant predictor-modifier interaction pairs are shown (p value for interaction<0.01).

OR, odds ratio; CI, confidence interval.

| | OR | 95% CI | p val |
|---|---|---|---|
| **Gender (ref=Female)** | | | |
| Male | 0.7 | 0.49-0.99 | 0.086 |
| **Ethnicity (ref=Caucasian)** | | | |
| South Asian | 0.39 | 0.2-0.77 | 0.018 |
| Afro-Carribean | 0.75 | 0.39-1.44 | 0.386 |
| Other | 0.58 | 0.25-1.33 | 0.245 |
| **Age (ref=18-55)** | | | |
| 55-64 | 3.54 | 2.1-5.99 | <0.001 |
| 65-74 | 6.72 | 4.01-11.25 | <0.001 |
| 75+ | 10.35 | 5.83-18.38 | <0.001 |
| **Body mass index** | | | |
| per 5 unit increase | 0.86 | 0.73-1.01 | 0.097 |
| **Smoking (ref=Never)** | | | |
| Past | 0.79 | 0.52-1.21 | 0.276 |
| Current | 0.52 | 0.33-0.8 | 0.01 |
| **Alcohol (ref=Never)** | | | |
| Past | 0.55 | 0.33-0.93 | 0.074 |
| Current | 0.95 | 0.63-1.44 | 0.824 |
| **Comorbidity (ref=No)** | | | |
| Diabetes | 1.63 | 1.12-2.39 | 0.023 |
| High cholesterol | 1.38 | 0.95-2.01 | 0.179 |
| Hypertension | 2.14 | 1.49-3.07 | <0.001 |
| Cardiovascular | 1.36 | 0.86-2.15 | 0.308 |
| Chronic respiratory | 0.87 | 0.54-1.39 | 0.805 |
| Chronic kidney | 0.83 | 0.41-1.67 | 0.712 |
| Chronic liver | 0.66 | 0.25-1.72 | 0.723 |
| Cancer | 1.56 | 0.87-2.81 | 0.275 |
| **Symptoms (ref=No)** | | | |
| Pain | 0.41 | 0.29-0.59 | <0.001 |
| Jaundice | 5.06 | 3.37-7.59 | <0.001 |
| Weight loss | 2.25 | 1.59-3.19 | <0.001 |
| Nausea | 0.58 | 0.4-0.84 | 0.008 |
| Vomit | 0.44 | 0.3-0.67 | <0.001 |
| Diarrhoea | 0.61 | 0.4-0.92 | 0.038 |
| Constipation | 0.8 | 0.52-1.22 | 0.583 |
| Fatigue | 2.18 | 1-4.77 | 0.1 |
| Loss of appetite | 1.05 | 0.43-2.57 | 0.917 |
| Pruritus | 4.34 | 1.21-15.6 | 0.049 |
| Steatorrhoea | 2.21 | 0.56-8.64 | 0.493 |
| Change in bowel habits | 1.38 | 0.91-2.08 | 0.237 |
| **Blood test (ref=Normal)** | | | |
| **Albumin** | | | |
| Low | 1.64 | 0.96-2.802 | 0.213 |
| High | 0.73 | 0.3-1.756 | 0.48 |
| **Alkaline phosphatase** | | | |
| Low | 0.34 | 0.15-0.77 | 0.015 |
| High | 3.3 | 1.86-5.863 | <0.001 |
| **Alanine aminotransferase** | | | |
| High | 2.54 | 1.6-4.041 | <0.001 |
| **Basophils** | | | |
| High | 1.06 | 0.72-1.547 | 0.772 |
| **Creatinine** | | | |
| Low | 1.83 | 0.93-3.619 | 0.239 |
| High | 0.99 | 0.44-2.222 | 0.979 |
| **Eosinophils** | | | |
| High | 0.85 | 0.45-1.588 | 0.772 |
| **Haemoglobin** | | | |
| Low | 1.85 | 1.29-2.658 | 0.003 |
| **Lymphocytes** | | | |
| Low | 2.03 | 1.3-3.182 | 0.008 |
| High | 1.73 | 0.18-16.93 | 0.629 |
| **Mean corpuscular volume** | | | |
| Low | 0.78 | 0.37-1.662 | 0.654 |
| High | 1.36 | 0.46-4.017 | 0.654 |
| **Neutrophils** | | | |
| Low | 0.57 | 0.08-3.964 | 0.564 |
| High | 1.32 | 0.86-2.022 | 0.516 |
| **Potassium** | | | |
| Low | 0.6 | 0.18-2.062 | 0.483 |
| High | 1.32 | 0.77-2.282 | 0.483 |
| **Platelets** | | | |
| Low | 0.58 | 0.21-1.599 | 0.33 |
| High | 2.21 | 1.07-4.594 | 0.1 |
| **Red blood cells** | | | |
| Low | 2.31 | 1.56-3.414 | <0.001 |
| **Sodium** | | | |
| Low | 4.1 | 1.69-9.966 | 0.006 |
| High | 0.41 | 0.12-1.361 | 0.214 |
| **Total bilirubin** | | | |
| High | 5.87 | 3.74-9.219 | <0.001 |
| **Urea** | | | |
| Low | 1.07 | 0.46-2.485 | 0.878 |
| High | 1.39 | 0.78-2.465 | 0.456 |
| **White blood cells** | | | |
| Low | 0.84 | 0.19-3.728 | 0.817 |
| High | 1.06 | 0.68-1.677 | 0.817 |

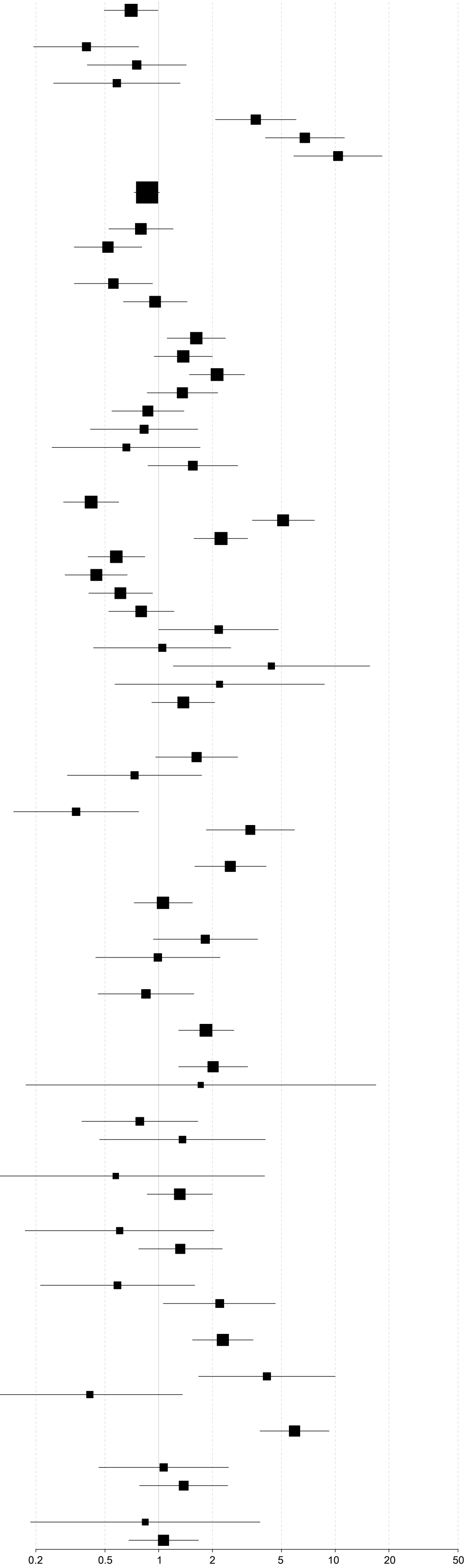

**Supplementary figure 1.** Forest plot showing crude association between study variables and odds of PDAC in comparison to non-malignant pancreatic disease group. The odds ratio (OR) and 95% confidence interval (CI) are derived from logistic regression model. The reported p values are corrected for multiple testing via Benjamini-Hochberg method.
